# Neutrophils cause critical illness in COVID-19 and reveal CDK6 inhibitors as potential treatment

**DOI:** 10.1101/2021.05.18.21256584

**Authors:** Hannes A. Baukmann, Justin L. Cope, Charles N. J. Ravarani, Colin Bannard, Margaretha R. J. Lamparter, Alexander R. E. C. Schwinges, Joern E. Klinger, Marco F. Schmidt

## Abstract

**Background:** Despite recent development of vaccines and monoclonal antibodies to prevent SARS-CoV-2 infection, treatment of critically ill COVID-19 patients remains an important goal. In principle, genome-wide association studies (GWAS) could shortcut the clinical evidence needed to repurpose drugs - the use of an existing drug for a new indication. However, it has been shown that the genes found in GWA studies usually do not encode an established drug target and the causal role for disease, a key requirement for drug efficacy, is unclear. We report here an alternative method for finding and testing causal target candidates for drug repurposing.

**Methods:** Rather than focusing on the genetics of the disease, we looked for disease-causing traits by searching for significant differences in 33 blood cell types, 30 blood biochemistries, and body mass index between an infectious disease phenotype and healthy controls. We then matched critically ill COVID-19 cases with controls that exhibited mild or no symptoms after SARS-CoV-2 infection in order to identify disease-causing traits by applying causal inference methods.

**Results:** We found high neutrophil cell count as a causal trait for the immune overreaction in critical illness due to COVID-19. Based on these findings, we identified the enzyme CDK6 as a potential drug target to prevent the immune overreaction in critical illness due to COVID-19.

**Conclusions:** The genetics of disease-causing traits turns out to be a rich reservoir for the identification of known drug targets. This is due to the usually larger datasets of traits, as they also cover healthy ones. A clinical trial testing CDK6 inhibitor palbociclib in critically ill COVID-19 patients is currently ongoing (ClinicalTrials.gov Identifier: NCT05371275).

## Introduction

The phenotype of critically ill coronavirus disease 2019 (COVID-19) status substantially differs from mild or moderate disease, even among hospitalized cases, by an uncontrolled overreaction of the host’s immune system^1–3^ – a so-called virus-induced immunopathology^4^ – resulting in acute respiratory distress syndrome (ARDS). The molecular mechanism leading to critical illness due to COVID-19 is still unclear. Identifying causal risk factors is central for prevention and treatment. Nonetheless, there is evidence that susceptibility and overreaction of the immune system to respiratory infections are both strongly heritable.^5,6^ A series of genome-wide association (GWA) studies have been conducted to investigate disease pathogenesis in order to find mechanistic targets for therapeutic development or drug repurposing.^7–10^ Treating the disease remains a top priority despite the recent development of vaccines preventing severe acute respiratory syndrome coronavirus 2 (SARS-CoV-2) infection due to the threat of new vaccine-resistant variants.

The results of 46 GWA studies comprising 46,562 COVID-19 patients from 19 countries have been combined in three meta-analyses by the COVID-19 Host Genetics Initiative.^10^ Overall, 15 independent genome-wide significant loci associations were reported for COVID-19 infection in general, of which six were found to be associated with critical illness due to COVID-19: 3p21.31 close to *CXCR6*, which plays a role in chemokine signaling, and *LZTFL1*, which has been implicated in lung cancer; 12q24.13 in a gene cluster that encodes antiviral restriction enzyme activators; 17q21.31, containing the *KANSL1* gene, which has been previously reported for reduced lung function; 19p13.3 within the gene that encodes dipeptidyl peptidase 9 (*DPP9*); 19p13.2 encoding tyrosine kinase 2 (*TYK2*); and 21q22.11 encoding the interferon receptor gene *IFNAR2*. The functions of the genes associated with these six loci are either related to host antiviral defense mechanisms or are mediators of inflammatory organ damage.

Nonetheless, using GWA data for drug development has several general drawbacks, which are particularly evident here with COVID-19. First, none of the reported genes encodes an established drug target. Rather, the exact function of the gene variants found in patients with critical illness due to COVID-19 is unclear. Therefore, it is questionable whether the gene product can be manipulated in function by a drug at all. Second, GWA studies only correlate genes with the disease. A causal relationship, which is important for drug development, cannot be deduced from this. Third, due to the currently limited sample size of GWA study datasets (<5,000 individuals), biologically relevant rare variants with small effect sizes cannot be detected.

Here, we present an approach for drug development or repurposing that is based on the genetics of disease-causing traits rather than the genetics of disease (Fig. 1). Using data from the UK Biobank^11^, critically ill COVID-19 patients were matched with a control group of COVID-19 patients with mild illness. Traits that differed significantly in cases and controls were further examined for causality with respect to critical illness in COVID-19 (Fig. 2). The genetics of these traits were further investigated to identify and test established target genes for drug repurposing. Focusing on the genetics of disease-causing traits reveals three advantages: First, disease-causing traits can more likely be manipulated with a drug via largely known druggable targets such as enzymes or receptors. Second, unlike a disease-associated gene, the function and, from there, causality of a gene for a trait is easier to verify. Third, the sample size of trait datasets is far greater than that of datasets specifically for COVID-19. For example, datasets on traits such as blood biochemistry often include >500,000 cases. Therefore, biologically relevant rare variants with small effect sizes can be detected.

**Fig. 1.**
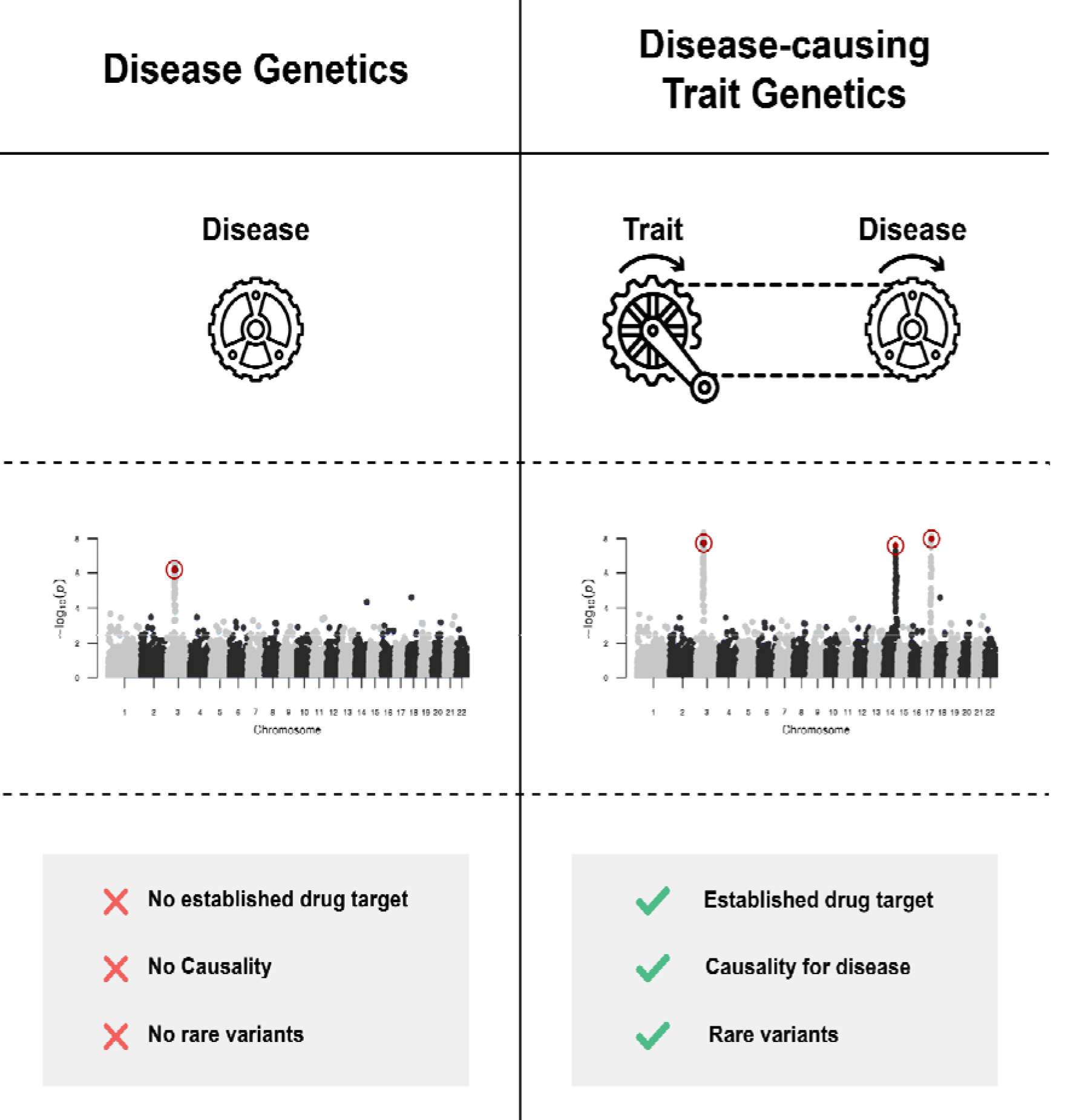
Disease genetics vs. disease-causing trait genetics for the identification of drug targets. Instead of focusing on disease genetics, genetics of disease-causing traits has three advantages: First, disease-causing traits are often more likely to be manipulated with a drug via largely known druggable targets such as enzymes or receptors. Second, unlike a disease-associated gene, the function and, from there, causality of a gene for a trait is easier to verify. Third, the sample size of trait datasets is far greater than that of datasets specifically for COVID-19.

**Fig. 2.**
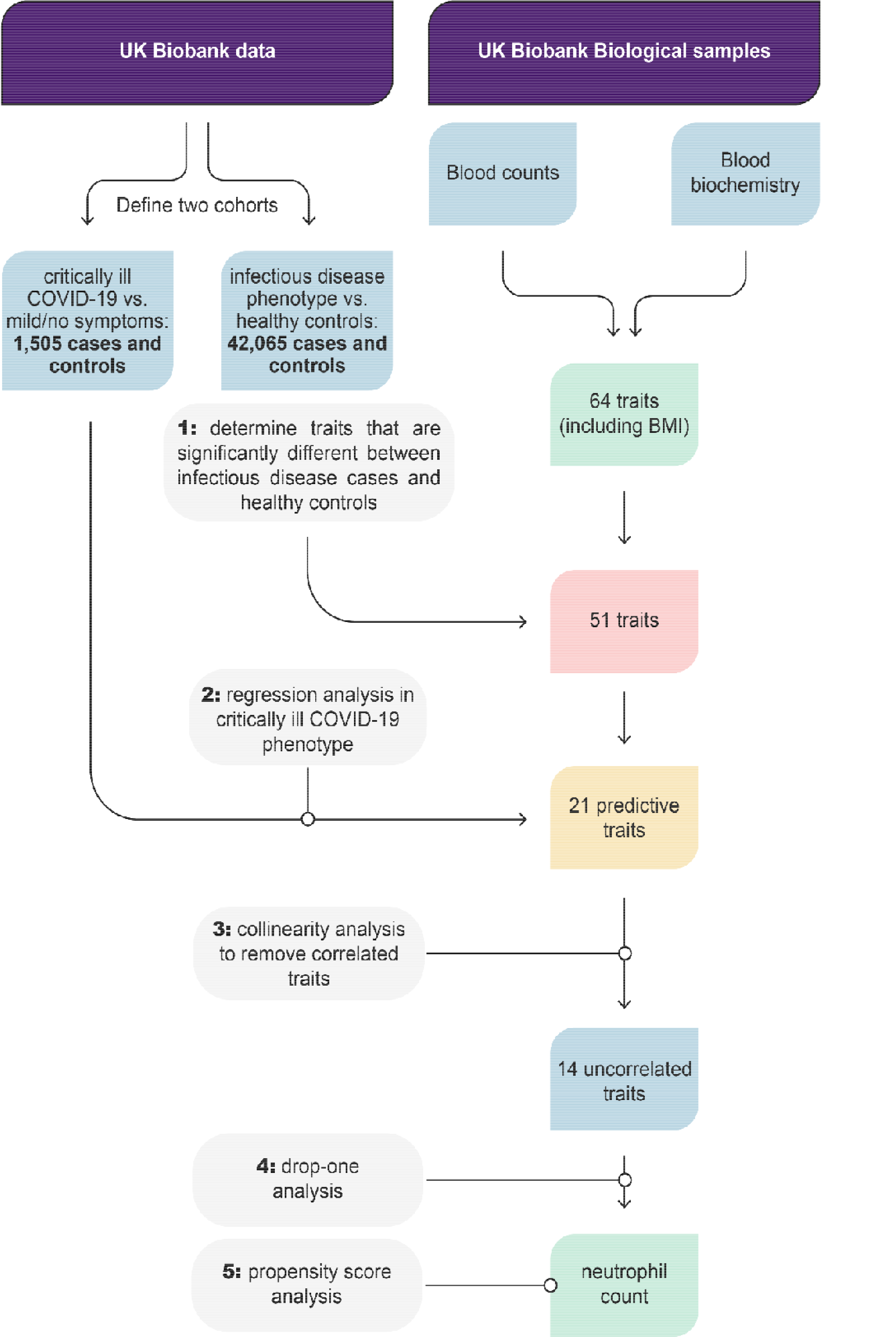
Workflow to identify traits leading to critical illness due to COVID-19. We identified significant differences in 64 candidate predictive traits between an infectious disease cohort and healthy controls. We used regression models to investigate the effect of these traits on critically ill COVID-19 cases compared to asymptomatic controls. Because highly dependent traits would not be significant in drop1 analysis, we first used collinearity testing to remove correlated traits. Using drop-one analysis, we identified neutrophil cell count as a trait that has a unique effect on critical illness in COVID-19.

**Fig. 3.**
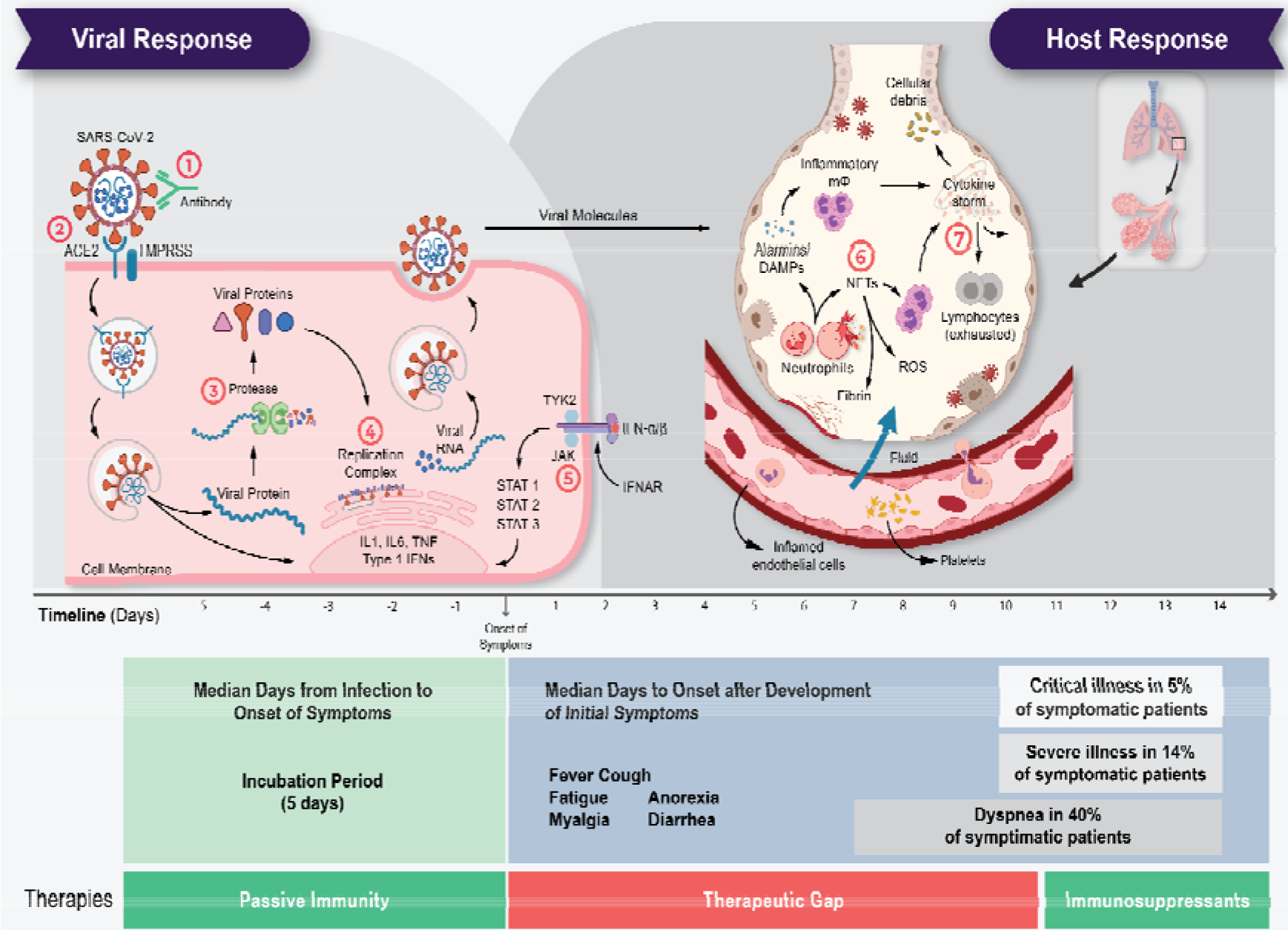
The life cycle of SARS-CoV-2 and the corresponding pathogenesis of COVID-19 display two phases: a viral response and a host-response phase. In the viral response phase, the virus enters the host cell and viral replication begins. Approximately five days after infection and successful replication, initial mild and moderate symptoms such as fever, cough, fatigue, anorexia, myalgia, and diarrhea are observed in conjunction with a decrease in lymphocyte cell count (lymphopenia). The following host-response phase determines the severity of the disease: in some patients, uncontrolled overreaction of the immune system – so-called virus-induced immunopathology – requires hospitalization and respiratory support due to acute respiratory distress syndrome (ARDS). Thus, severe cases of COVID-19 originate from an immune overreaction rather than from the viral infection itself. Currently, there are seven drug mechanisms described: ① Passive immunity; ② Entry inhibitors; ③ Protease inhibitors; ④ Polymerase inhibitors; ⑤ JAK inhibitors; ⑥ NETosis inhibitors; ⑦ Immunosuppressants.

## Methods

### Recruitment of cases and controls

We downloaded the rich information made available by the UK Biobank project on October 25, 2021. COVID-19 test results up until 18^th^ October 2021 were collected, and cases were defined as reported previously.^8^

Briefly, 1,505 severe cases were defined as patients who died or were hospitalized due to COVID-19 (cause of death or diagnosis containing ICD10 codes U07.1 or U07.2) or were ventilated (operation codes E85.*) in 2020 or 2021 and tested positive for SARS-CoV-2 infection. Individuals that were tested positive for SARS-CoV-2, but did not die or were critical due to COVID-19 and were not ventilated, were defined as potential mild COVID-19 controls.

The infectious disease phenotype was created based on UK Biobank data for respiratory infections, acute respiratory distress syndrome (ARDS), influenza, and pneumonia with hospitalization or death as a result. We aggregated hospital in-patient and death register data for ICD codes corresponding to J00-J06 (“Acute upper respiratory infections”), J09-J18 (“Influenza and pneumonia”), J20-J22 (“Other acute lower respiratory infections”), and J80 (ARDS), yielding 42,065 cases. The remaining individuals from the UK Biobank were defined as potential healthy controls.

For both cohorts, cases and controls were filtered for European ancestry (“British”,“Irish”, and “Any other white background”), and individuals with missing age and sex information were discarded. For both cohorts, controls were then matched to the same number of cases based on age and sex.

Variants reported by Pairo-Castineira *et al*.^*8*^ and Ellinghaus *et al*.^*7*^ as well as variants reported by the ClinVar database^12^ for the genes reported by the papers were included in the dataset.

### Screening for significant traits

The UK Biobank contains data on biological samples taken years before potential infection upon registration of individuals to the program, including 33 blood cell counts and 30 blood biochemistry measurements, and body mass index. In order to identify traits that are significantly different between the infectious disease cohort and healthy controls, we performed either independent two-sample t-test or Wilcox rank sum test from the R package stats (https://www.rdocumentation.org/packages/stats/versions/3.6.2), depending if the trait follows a normal distribution or not. We applied a Bonferroni-corrected *p*-value threshold of p < α/n = 0.05/64. In five instances, the *p*-values were too small to be represented properly, and were instead set to 1.0E-297.

### Regression modeling

Logistic regression models were fitted using the *glm* function in R (www.R-project.org).

### Collinearity testing

We applied a collinearity threshold of 0.5 and subset from the data the trait pairs where the absolute collinearity estimate is greater or equal to the collinearity threshold. We then iteratively removed the trait with the lower regression coefficient of that pair.

### Drop-one analysis

A drop-one model comparison procedure was performed using the *drop1()* function in R (www.R-project.org) in order to determine whether each of a set of traits accounts for unique variance in critically ill COVID-19 disease status. The formula of BMI + high light scatter reticulocyte count + erythrocyte distribution width + neutrophil count + lymphocyte count + alkaline phosphatase + apolipoprotein A + C-reactive protein + cystatin C + gamma glutamyltransferase + glucose + SHBG + triglycerides + vitamin D was used to predict critical illness due to COVID-19. Single terms were deleted and the F value is calculated to perform an F-test to derive the Pr(>F) value, where low values indicate that a model that does not include this term is significantly different from the full model.

### Propensity score analysis

Using the method of Imai and Van Dyk^13^, individuals are split into deciles who have a similar propensity for a treatment (neutrophil count) given the covariates (the risk factors age, sex, BMI, C-reactive protein, cystatin C (as a proxy for cardiovascular disease), alanine aminotransferase (as a proxy for chronic liver disease), and creatinine (as a proxy for chronic kidney disease)). We then estimated the effect of treatment on severe COVID-19 within each of the groups. The effect across these groups is examined and the average effect of treatment is calculated over the groups to give an estimate of effect of treatment independent of the covariates.

### GWA analysis

Prior to genome-wide association analysis, we took steps to remove biases by submitting UK Biobank genotypes to a series of quality control steps using PLINK 2.0^14^. First, we extracted variants on autosomal chromosomes. Then, we filtered samples for European ancestry, and we further dropped all samples with missing data for the phenotype of interest (neutrophil cell count) or for any of the following covariates: age, sex, BMI, and genetic principal components. These initial filtering steps left us with 444,114samples and 784,256 variants. Next, we filtered variants for minor allele frequency (MAF) using a threshold of 0.01 for the aggregate frequency and count of non-major alleles, since extremely rare alleles may indicate genotyping errors and furthermore are cases where power for detecting variant-to-phenotype associations is lacking. We then filtered variants based on missingness in the dataset with a threshold of 0.1, excluding variants where genotyping information is unavailable or of poor quality for more than 10 percent of subjects. Next, as an additional guard against genotyping errors, variants deviating from Hardy-Weinberg equilibrium were removed where exact test *p*-values fell below the threshold of 1e-15. We then filtered samples with a missingness threshold of 0.1, excluding samples where genotyping information is unavailable or of poor quality for more than 10 percent of variants. This yielded a final dataset with a total of 444,109 samples and of 509,485 quality controlled variants. Finally, a genome-wide association analysis was performed in two steps with REGENIE^15^. In the first step, a whole genome regression model was fitted using ridge regression, and in the second step, variants were tested for association with the continuous neutrophil cell count phenotype conditioned on the prediction of the model from the prior step using the “leave one chromosome out” scheme (LOCO) to avoid proximal contamination. In both steps, the first four genetic principal components were included as covariates.

### Statistical power calculation

The calculation of the effect size required to achieve a certain statistical power based on a fixed *p*-value threshold is based on https://www.mv.helsinki.fi/home/mjxpirin/GWAS_course/material/GWAS3.html. Due to the high computational cost, a random slice of 10% of the variants from the GWAs was used in these calculations.

### Mendelian randomization

We used independent GWAS summary data for neutrophil cell count (exposure) published by Vuckovic *et al*.^*16*^ (GCST90002398 downloaded January 15th 2021) and summary data for critically ill COVID-19 status (outcome) published by the COVID-19 Host Genetics Initiative (https://www.covid19hg.org/results - COVID19hg GWAS meta-analyses round 5 release date January 18th 2021). Two-sample MR analyses were done as previously described.^10^

## Results

### Screening for traits associated with infectious disease

Using UK Biobank data^11^, we identified 42,065 individuals with respiratory infections, acute respiratory distress syndrome (ARDS), influenza and pneumonia, which serve as our infectious disease cohort. In order to explore how the infectious disease cohort differs from healthy controls, we screened 64 candidate predictive traits that had been measured years before the individuals were affected. We observed Bonferroni-corrected statistically significant differences (*p* < □/*n* = 0.05/64)^17^ in 51 traits confirmed by independent two-sample t-test and two-sided Wilcox rank sum test (Fig. 2 and SI Fig. 1).

### Regression modeling

Furthermore, we identified 1,505 patients who were hospitalized due to SARS-CoV-2 infection and who required respiratory support and/or died due to infection.^18^ These patients were defined as cases and matched to controls that were infected with SARS-CoV-2, but showed no and only mild symptoms. Carrying over the 51 traits identified in the previous step, we used regression modeling to investigate the effect of these traits on critically ill COVID-19 status. Out of the 51 traits, 21 traits were significant predictors of critical illness due to COVID-19 with a Bonferroni-corrected significance threshold of *p* < □/*n* = 0.05/51 (Fig. 2 and SI Tab. 1).

### Collinearity analysis

Collinearity is the correlation between predictor variables in a regression model. Therefore, collinearity between traits would impact the results of the drop-one analysis. We first identified traits to remove in order to solve this issue. Seven traits were thus excluded from further analysis: Leukocyte count, reticulocyte count, reticulocyte percentage, high light scatter reticulocyte percentage, immature reticulocyte fraction, HDL cholesterol, and glycated hemoglobin (HbA1c) (Fig. 2 and SI Tab. 2).

### Drop-one analysis

The drop-one analysis compares all possible models that can be constructed by dropping a single model term and evaluating its impact on the regression model. The drop-one analysis revealed that only neutrophil count explains unique variance in critically ill COVID-19 status to a Bonferroni-corrected significance threshold of *p* < □/*n* = 0.05/14 (Fig. 2 and SI Tab. 3).

### Propensity score analysis

Propensity score analysis is a technique for estimating the effect of a treatment on an outcome independent of covariates. We employed propensity score stratification using the propensity function of Imai and van Dyk^13^ in order to estimate the effect of the treatment on critical illness in COVID-19 independent of the known risk factors for critical illness in COVID-19: age, sex, BMI, C-reactive protein (as a proxy for autoimmune disease), cystatin C (as a proxy for cardiovascular disease), alanine aminotransferase (as a proxy for chronic liver disease), and creatinine (as a proxy for chronic kidney disease). Neutrophil count has in fact a significant effect on critical illness in COVID-19 (*p* = 1.8228E-06, estimated effect = 0.13177±0.028456) (Fig. 2 and SI Tab. 4).

### Trait genetics analysis

We next focused on the genetics of neutrophil cell number. We performed a GWA analysis on neutrophil cell count using the entire UK Biobank (471,532 participants) (SI Fig. 2). We compared our results with previously published statistics from the NHGRI-EBI GWAS catalog^19^ and were able to confirm them. Subsequently, gene variants were analyzed for agents associating the ChEMBL database^20^ with the associated genes with a significance of -log *p* = 80 or better (SI Tab. 5). Since no clear drug-to-gene assignment was possible for the gene variants of the HLA haplotype on chromosome 6, we focused on all other significant gene variants in the following. The most significant gene variants were found in the *PSMD3* gene and are associated with bortezomib and carfilzomib according to the ChEMBL database. This is followed by gene variants in the *CDK6* gene, that is associated with the drugs palbociclib, ribociclib, fulvestrant, abemaciclib, trilaciclib, apremilast and dexamethasone. Furthermore, gene variants and associated drugs were found in the genes *NR1D1* (lithium), *THRA* (levothyroxine, liothyronine, aspirin, and lithium), *CXCR2* (clotrimazole, acetylcysteine, and ibuprofen), as well as *PLAUR* (filgrastim and ruxolitinib). Bortezomib and carfilzomib are proteasome inhibitors approved for cancer therapy, whereas *PSMD3* encodes one of the non-ATPase subunits of the 19S regulatory lid. Therefore, bortezomib and carfilzomib do not bind directly to the protein of the *PSMD3* gene. In contrast, the drugs already approved for breast cancer abemaciclib, ribociclib, trilaciclib, and palbociclib bind directly to the protein encoded by the *CDK6* gene, cyclin-dependent kinase 6 (CDK6). Because of the high significance and direct binding of the drugs to CDK6, we considered this to be the best potential target for lowering neutrophil counts, and thus, preventing immune system overreaction in critical illness due to COVID-19 in high-risk individuals with high neutrophil counts.

We also conducted GWA analyses with the cases and controls defined earlier and a random population of the same size (n = 3,010) (SI Figs. 3 and 4, respectively). In both cases, we could not identify variants with genome-wide significance. Statistical power analysis shows that this is due to the lack of statistical power in GWA analyses with that few individuals (SI Fig. 5).

### Genetic proxy regression modeling of CDK6 inhibitor treatment

We then used 58 reported variants in the *CDK6* gene to predict neutrophil count in all subjects of European ancestry (n = 471,532), the cases and controls defined earlier (n = 3,010) and a random population of the same size (n = 3,010) as a genetic proxy of a CDK6 inhibitor treatment. While most of the variants had a significant effect on neutrophil count in the whole population, none of the variants showed a significant effect in the other two sets of individuals (SI Tab. 6). This suggests that the case/control data set is too small to detect the effect of *CDK6* variants on neutrophil count.

### Mendelian randomization

Mendelian randomization (MR) is a robust and accessible tool to examine the causal relationship between an exposure variable and an outcome from GWAS summary statistics.^21^ We employed two-sample summary data Mendelian randomization to further validate causal effects of neutrophil cell count genes on the outcome of critical illness due to COVID-19. We used independent GWAS summary data for neutrophil cell count (exposure) published by Vuckovic *et al*.^*16*^ and summary data for critical illness in COVID-19 (outcome) published by the COVID-19 Host Genetics Initiative^10^. As shown in the Supplementary Information Tab. 4, instrumental variable weight (IVW) was significant with a *p*-value of 0.01199 when we used a lenient clumping parameter of r = 0.2 and 1,581 SNPs whereas we observed no significant IVW when we used strict clumping parameters of r = 0.01 and 567 SNPs (SI Tab. 7).

## Discussion

In classical GWA studies, drug targets are rarely found. That is because GWA hits correlate with disease, but their causality, which is compelling for drug development, is not proven. Moreover, rare variants with small effect sizes are not found because of sample sizes that are drastically limited by the number of patients available for study. In contrast, here we describe a method that prioritizes the identification of traits with a causal role in disease pathogenesis. Subsequent investigation of the genetics of the disease-causing traits enables the discovery of drug targets that would not be found in classical GWA studies because of typically small sample sizes.

Our approach was as follows. First, we identified significant differences in 64 predictive characteristics between a cohort of infectious disease and healthy control subjects from the UK Biobank. Using regression models, we examined the effects of these characteristics on severely ill COVID-19 cases compared with mild control cases. Because highly correlated characteristics would be missed in a drop-one analysis, collinear (non-independent) characteristics were first removed. Of the remaining characteristics, neutrophil count was identified as a characteristic that had a unique impact on critical illness in COVID-19 independent of other characteristics. Age, male gender, obesity, type 2 diabetes, cardiovascular disease, chronic liver and kidney disease have been previously described as risk factors for the severe course of COVID-19.^22^ Based on the characteristics measured in the UK Biobank, we used these risk factors or surrogate factors as confounders in the propensity score analysis. Finally, the propensity score analysis confirmed the causal effect of neutrophil count on severe COVID-19 independent of these risk factors.

The role of neutrophil cell count in COVID-19 can be explained by the previously reported disease mechanism.^23^ Neutrophils are white blood cells and an important component of our host defense against invading pathogens. Critical illness in COVID-19 is characterized by infiltration of the lungs with macrophages and neutrophils that cause diffuse lung alveolar damage, the histological equivalent to ARDS (Fig. 23).^22,24,25^ Neutrophils develop so-called neutrophil extracellular traps (NETs), web-like structures of nucleic acids wrapped with histones that detain viral particles, through NETosis, a regulated form of neutrophil cell death.^26^ However, ineffective clearance and regulation of NETs result in pathological effects such as thromboinflammation.^27^

Ultimately, we focused on the genetics of neutrophils and came across the *CDK6* gene. *CDK6* encodes cyclin-dependent kinase 6, an enzyme involved in cell division, for which three drugs have already been developed and approved for the treatment of breast cancer. To better understand the role of CDK6 in neutrophil count, we defined the SNP rs445, which is known in the literature, as a genetic proxy for treatment with CDK6 inhibitors. To do this, we use regression models with rs445 as a variable to predict neutrophil cell count in the three different datasets: the full UK Biobank (471,532 cases), our case-control dataset comparing severe COVID-19 progression versus mild progression (3,010), and a randomly selected cohort from the UK Biobank with the same sample size of 3,010 cases. Only in the cohort from the entire UK Biobank did we detect an effect of rs445 on neutrophil count. The effect size of rs445 on neutrophil cell count is too small for rs445 to show significant effects in smaller data sets. That is supported by our statistical power analysis we conducted with the three datasets (SI Fig. 5).

Cyclin-dependent kinases (CDK) 4 and 6 have been previously described as regulators of NETosis. CDK4/6 inhibitors block NETs formation in a dose-responsive manner but do not impair oxidative burst, phagocytosis, or degranulation.^28^ This indicates that CDK4/6 inhibition specifically affects NET production rather than universally modulating inflammatory pathways (in contrast to immunosuppressants such as dexamethasone or interleukin-6 inhibitors). This is supported by Grinshpun *et al*.’s report that COVID-19 progression was halted for a breast cancer patient on CDK4/6 inhibitor therapy. Once the drug was withdrawn, the full classic spectrum of illness appeared, including oxygen desaturation necessitating a prolonged hospital stay for close monitoring of the need for invasive ventilations.^29^ Selective inhibition of NETosis is a particularly attractive treatment because CDK4/6 inhibitors can prevent the cytokine storm and, thus, later intensive care.

In parallel, we performed Mendelian randomization (MR) with neutrophil count as exposure and critically ill COVID-19 course as outcome. The literature describes either no effect^30^ or a slightly negative association^31^ for this scenario. In our experiments, we saw the same result depending on how strictly clumping parameters were selected according to linkage disequilibrium (LD). If clumping was strict, we saw no effect. When we selected more variables due to a less stringent LD threshold, we found that a higher number of neutrophil cells seems to protect against the critical illness in COVID-19. However, the role of neutrophils as a driver of critical illness due to COVID-19 has been clearly described in the literature (see above). Why do we get this result in MR that is contrary to clinical observation? The reason can be explained by sample size in a manner analogous to the discussion of our regression analyses with rs445. As our statistical power analysis has shown, large sample sizes are needed to obtain a large number of gene variants with strong effect sizes. MR only works if a sufficient number of gene variants (instrument variables) with strong effect sizes for exposure and outcome are available. The summary statistics of neutrophil cell count and severe COVID-19 progression underlying MR show an imbalance of sample sizes. The here used statistics of the neutrophil cell count are based on 408,112 cases, whereas the statistics of critical illness in COVID-19 are based on only 5,582 cases. Ultimately, this leads to insufficient overlap of variables with the necessary effect size to generate a signal in Mendelian randomization. The artificial extension of the overlap by a less strict LD threshold seems to favor the amplification of false signals.

In conclusion, identifying drug targets from GWA data is challenging because of sample sizes limited by patient numbers and the accompanying high-dimensionality of the data structure. In addition, GWA studies only reflect associations and do not provide information on causality. In contrast, we have developed a workflow that enables the identification of causal drug targets via the identification and investigation of disease-causing traits. By focusing on the genetics of disease-causing traits, we can leverage larger sample sizes to reveal rare gene variants with small effect sizes. We applied our workflow to critical illness in COVID-19. We identified neutrophils as causal drivers of the disease. In addition, we found CDK6 as a drug target to reposition the already approved breast cancer drug palbociclib for potential preventive treatment of COVID-19. In the case reported by Grinshpun *et al*.,^29^ the CDK4/6 inhibitor was administered prior to infection, therefore it was not harmful in the early course of the disease (like immunosuppressants^32^), but protected against thromboinflammation and thus prevented the necessity of intensive care. Another advantage rendering CDK6 an attractive drug target is that since it is a human protein, mutations of the virus do not influence drug action – in stark contrast to vaccines and antivirals. Ultimately, CDK4/6 inhibitors could be used against all virus-induced immune pathologies, and thus also contain future pandemics of novel viruses. A clinical trial testing a CDK6 inhibitor in critically ill COVID-19 patients is currently ongoing.

## Data Availability

Following publication of major outputs all anonymised data will be made available on reasonable request to the corresponding author providing this meets local ethical and research governance criteria.

## Acknowledgment

The research has been conducted using the UK Biobank Resource under Application no. 36226. We thank Radi Hilaneh for making Fig. 1, Fig. 2, and Fig. 3. The research work was supported by the *Investitionsbank des Landes Brandenburg* (ILB), the European Regional Development Fund (ERDF), and the European Social Fund+ (ESF+). Access to the UK Biobank was funded by the EIT Health Digital Sandbox program to access European biobank data (grant number 2019-DS1001-3754). We also thank the program ‘digital solutions made in Brandenburg’ (digisolBB) for its continued support.

## Competing interests

H.A.B., J.L.C., C.N.J.R., M.R.J.L., J.E.K., and M.F.S are employees of biotx.ai GmbH. A.R.E.S was an employee of biotx.ai GmbH.

## Supplementary information

**SI Fig. 1.**
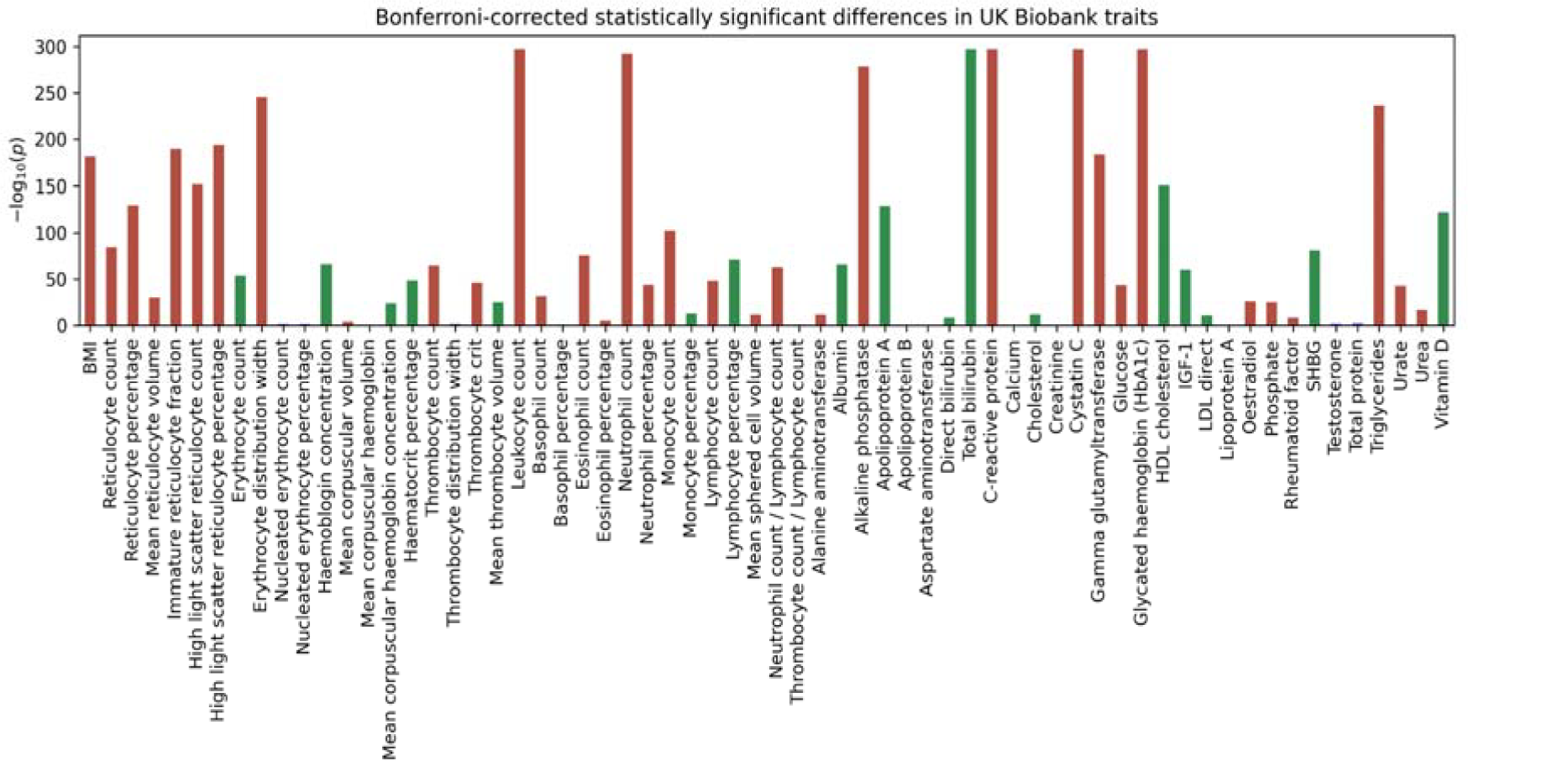
Bonferroni-corrected statistically significant differences in 64 traits identified using independent two-sample t-test and Mann-Whitney U test. Red and green columns indicate traits that are significantly increased in infectious disease cases or healthy controls, respectively.

**SI Fig. 2.**
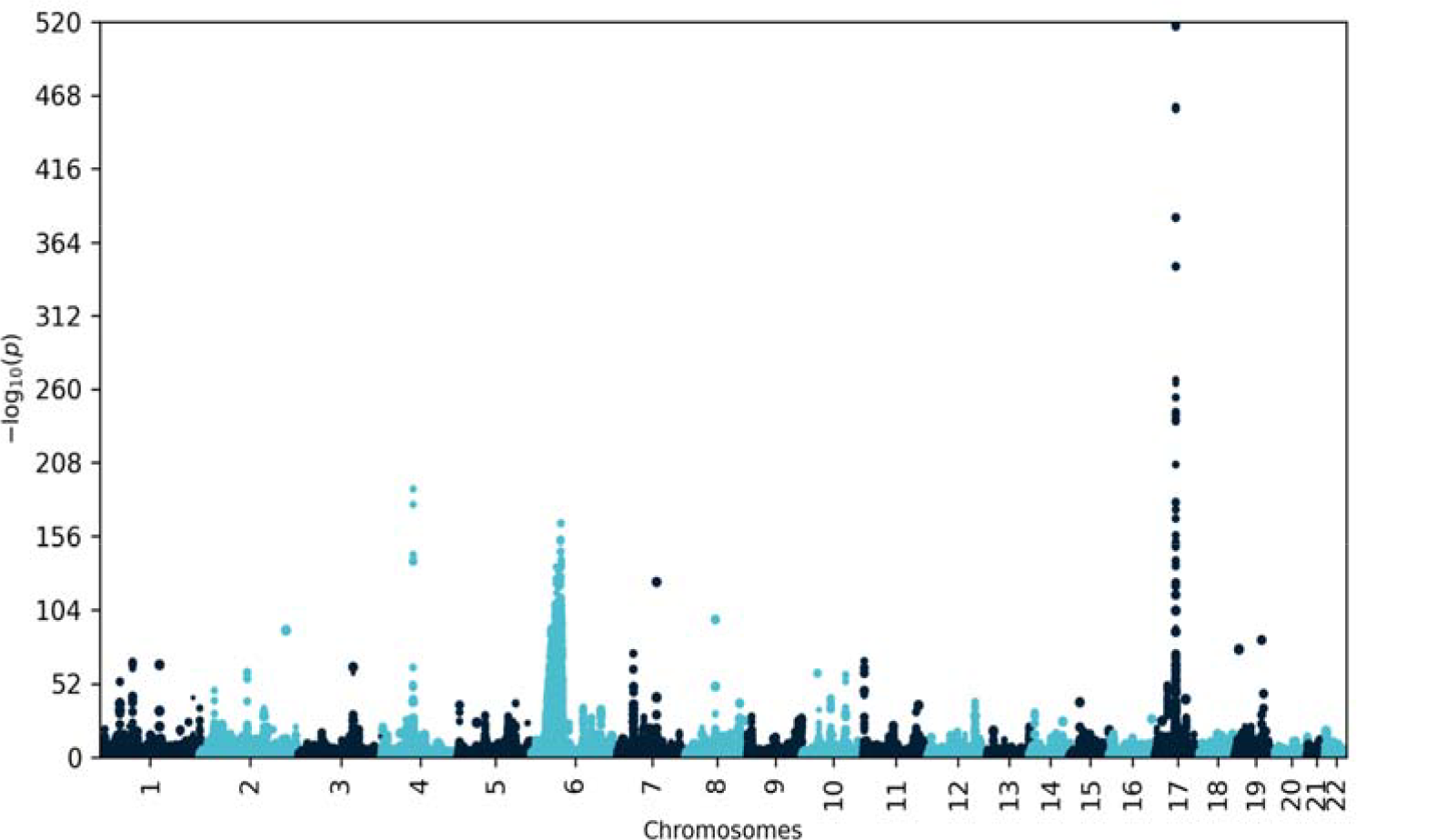
Manhattan plot of neutrophil cell count (n = 471,532). The significantly associated variants in the *CDK6* gene (-log *p*-values in parentheses) are rs445 (123.637), rs2282989 (42.226), rs42041 (30,014), rs42030 (20.325), and rs78366656 (15.206) on chromosome 7.

**SI Fig. 3.**
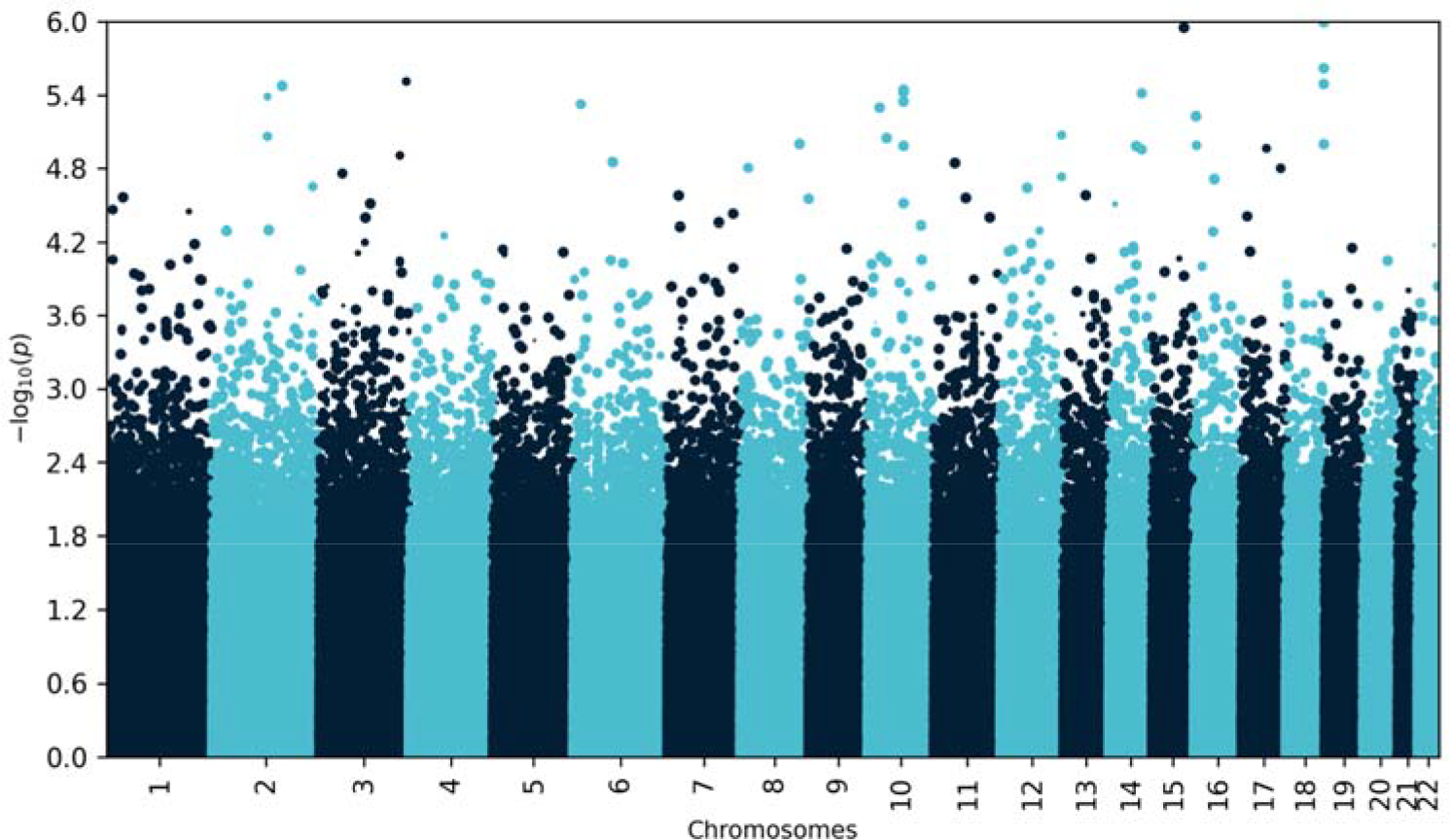
Manhattan plot of neutrophil cell count using only cases and controls (n = 3,010).

**SI Fig. 4.**
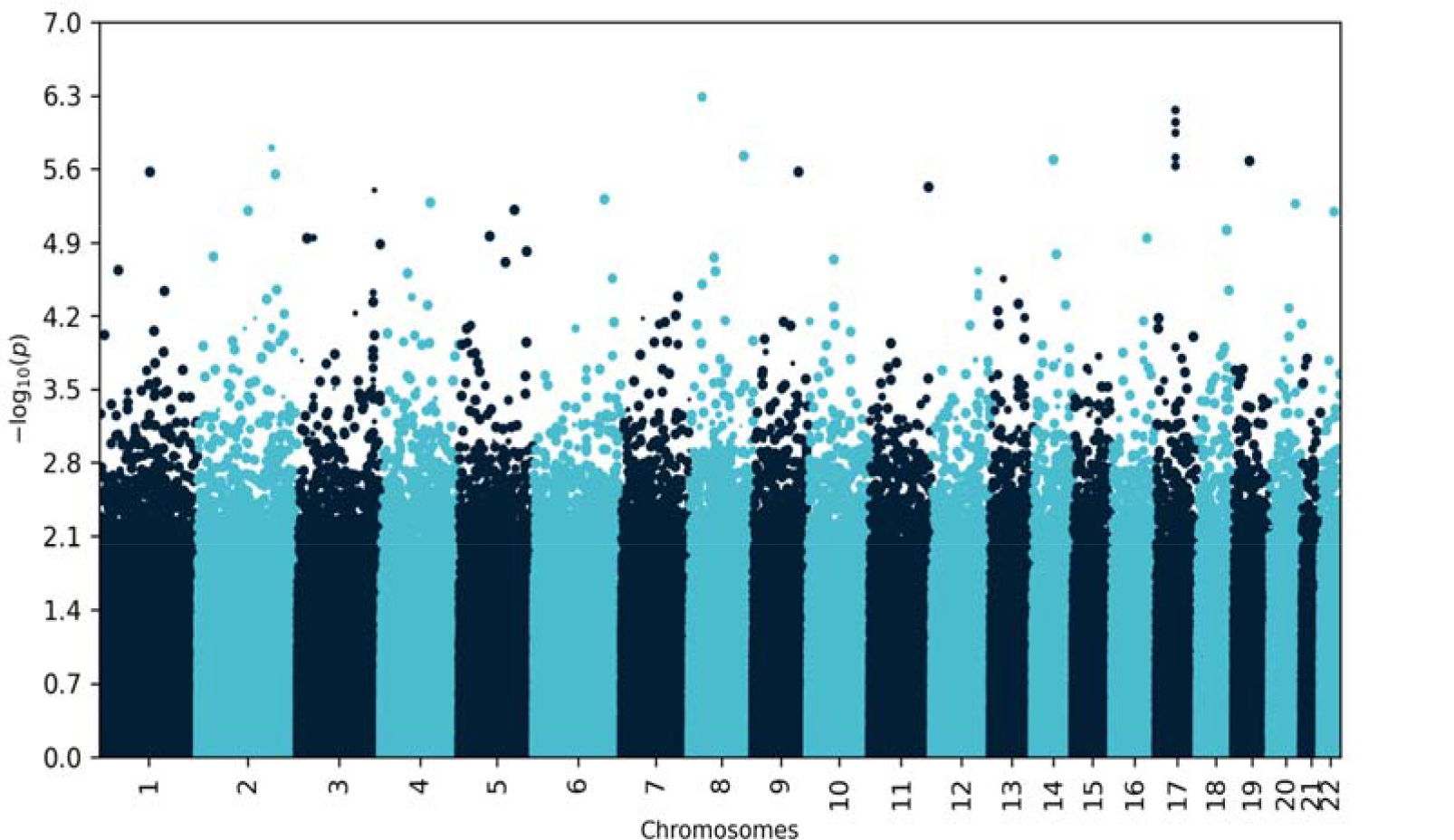
Manhattan plot of neutrophil cell count using a random subset of the filtered population (n = 3,010).

**SI Fig. 5.**
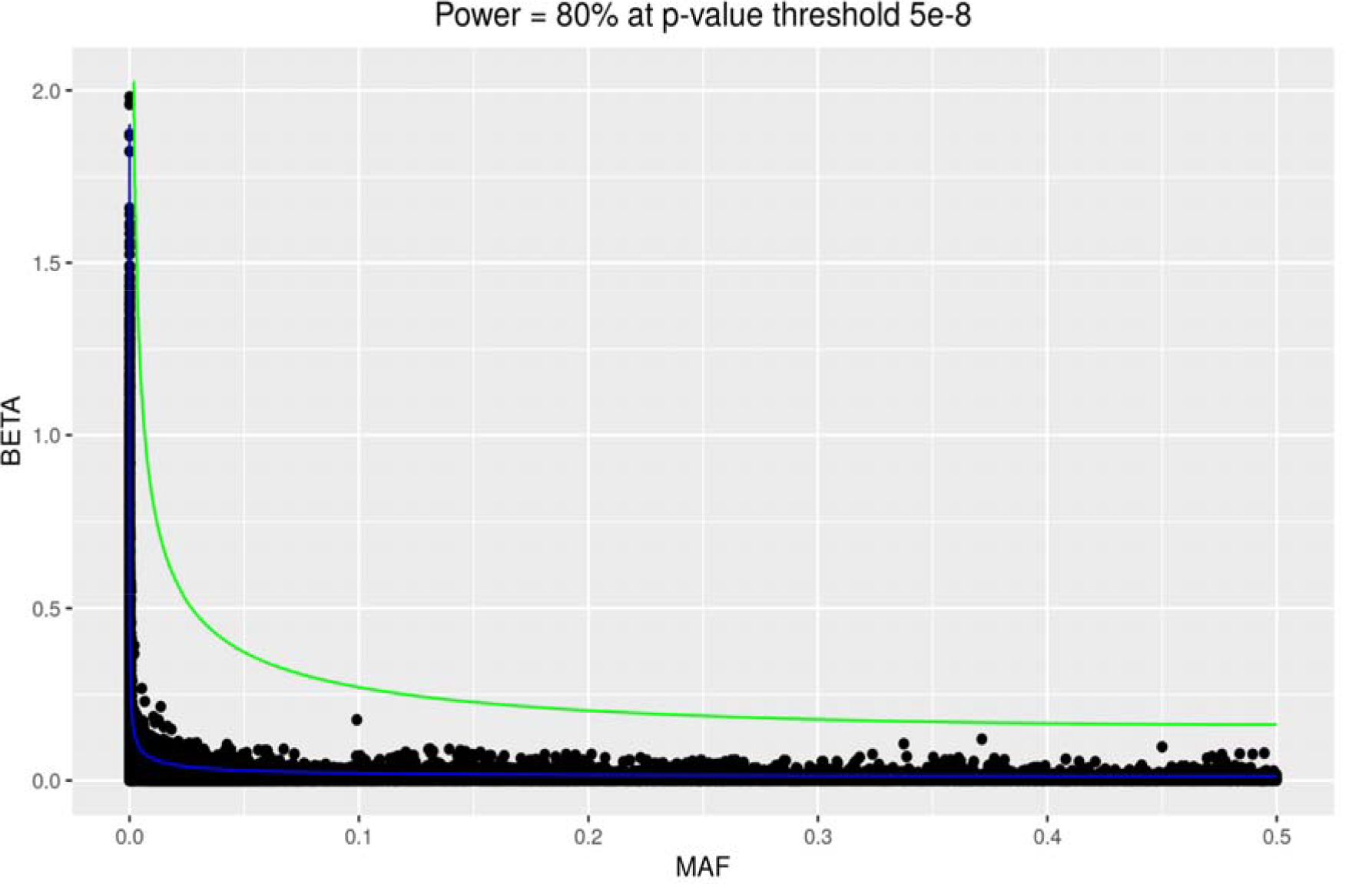
Statistical power calculations. Dots represent variants and their effect size (beta, log Odds Ratio) for neutrophil count as determined by the GWAS (n = 471,532). The lines represent the effect size required to achieve a statistical power of 80% at a *p*-value threshold of 1e-15 in the full GWAS (blue) and the GWAS with a random subset of the same size and the cases and controls for severe COVID-19 (green) (n= 3,010).

**SI Tab. 1.**
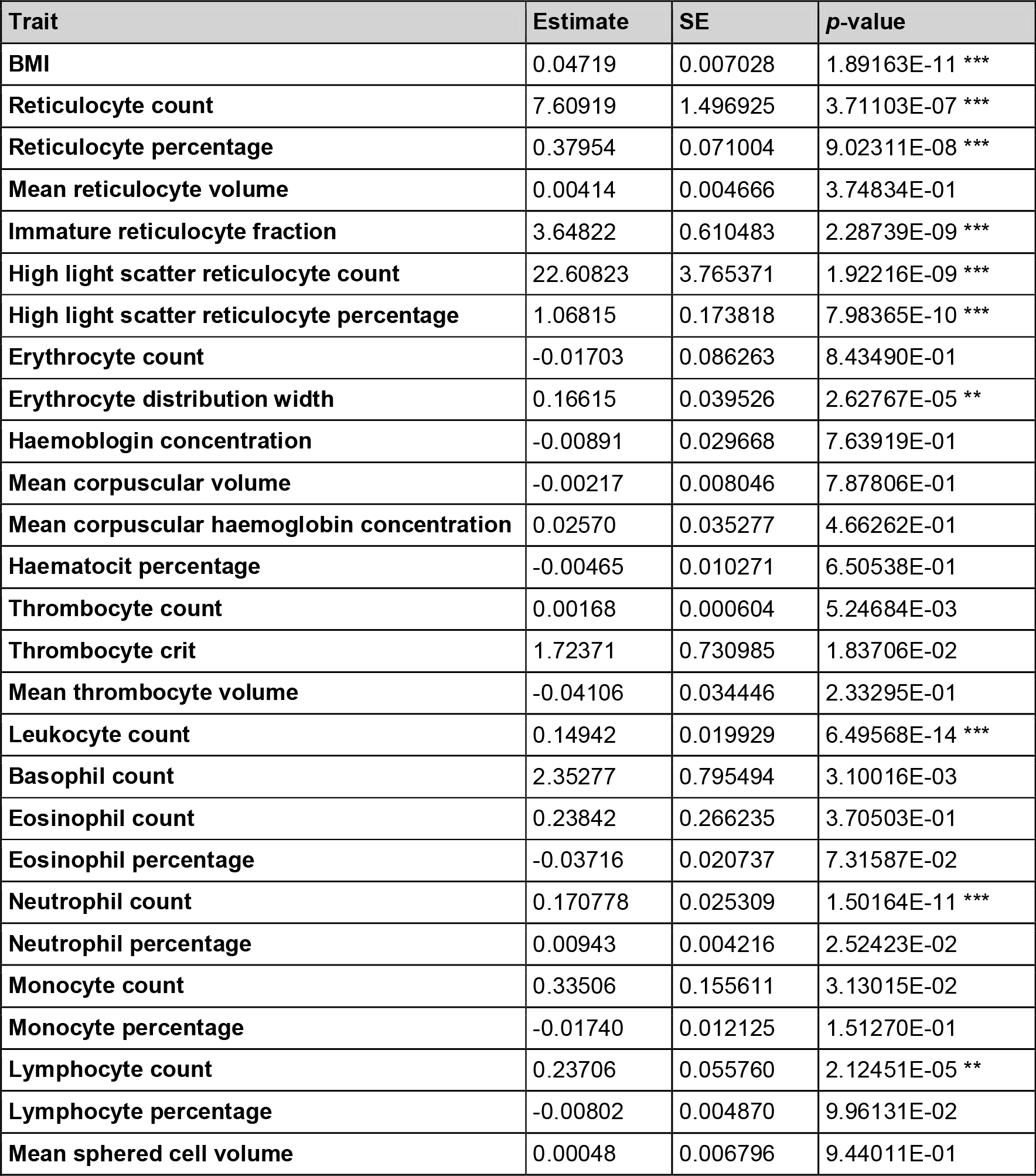
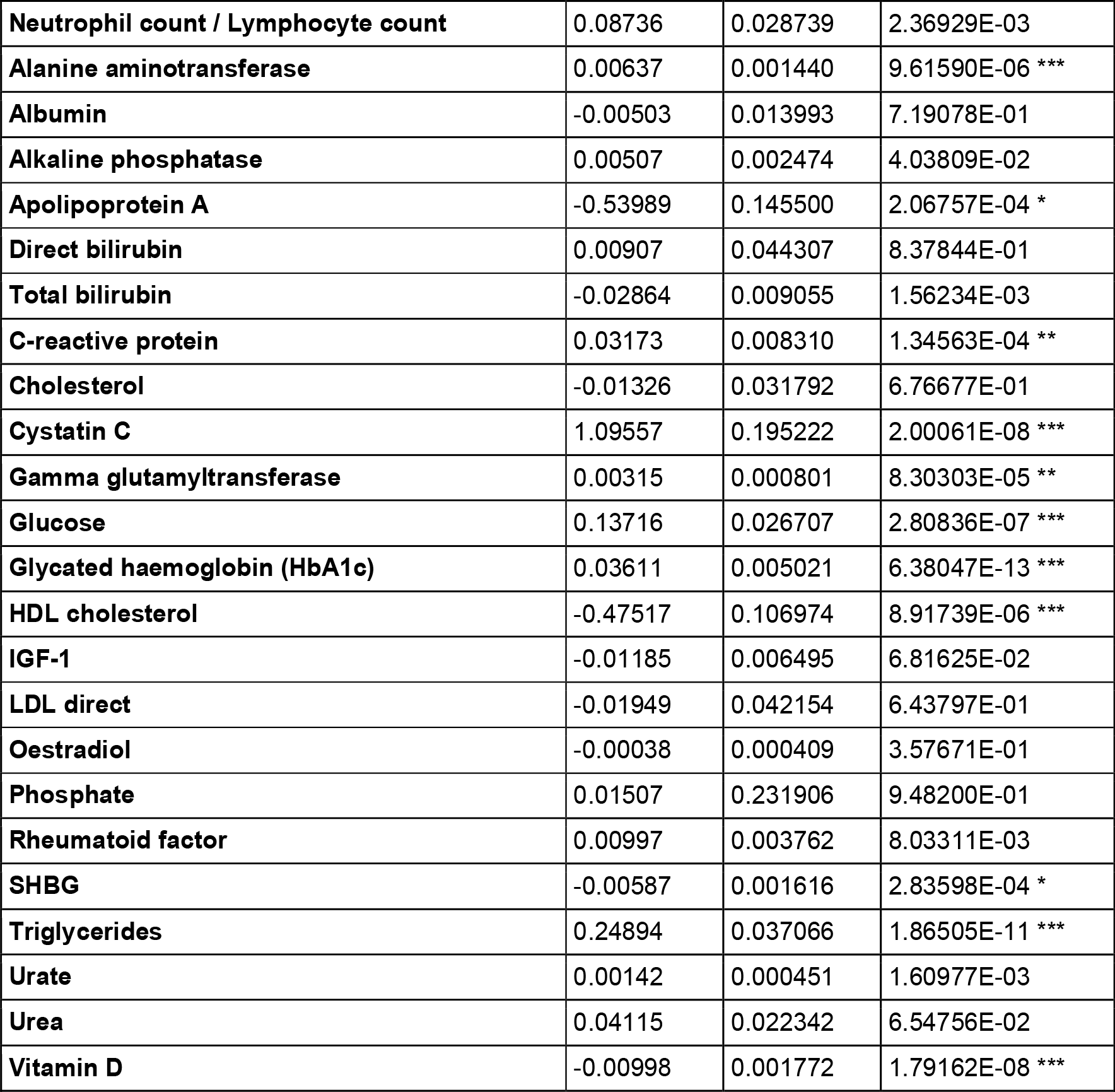
Critical illness in COVID-19 was regressed on the traits significantly different between infectious disease cases and healthy controls. Significance thresholds are indicated by asterisks, where three asterisks indicate *p*-values below 0.001/51, two indicate *p*-values below 0.01/51, and one asterisk indicates *p*-values below 0.05/51.

**SI Tab. 2.**
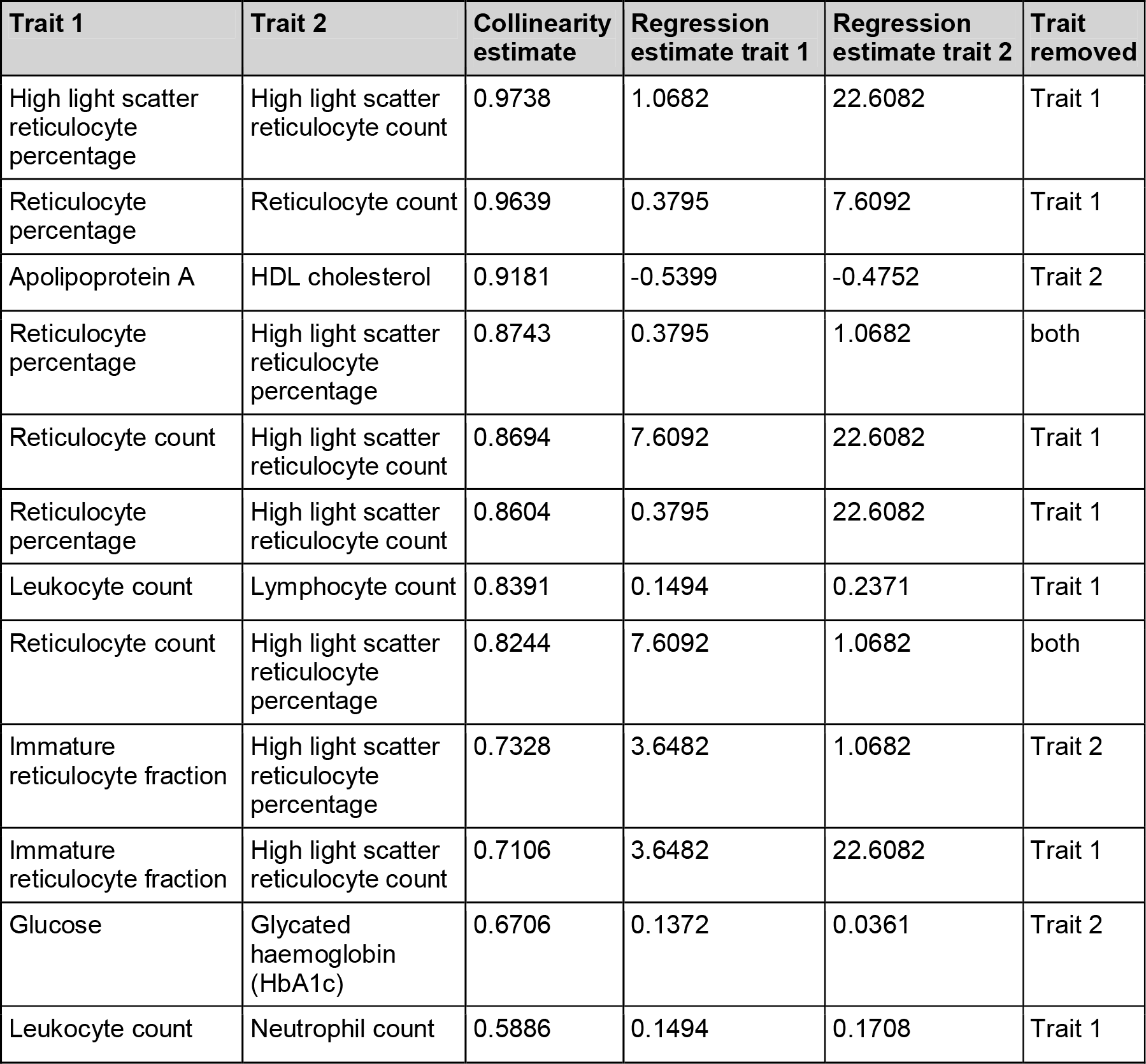
Collinearity estimates greater than 0.5 between the 21 traits significant in regression analysis. The traits with the lower regression estimates are removed.

| Trait 1 | Trait 2 | Collinearity estimate | Regression estimate trait 1 | Regression estimate trait 2 | Trait removed |
| --- | --- | --- | --- | --- | --- |
| High light scatter reticulocyte percentage | High light scatter reticulocyte count | 0.9738 | 1.0682 | 22.6082 | Trait 1 |
| Reticulocyte percentage | Reticulocyte count | 0.9639 | 0.3795 | 7.6092 | Trait 1 |
| Apolipoprotein A | HDL cholesterol | 0.9181 | -0.5399 | -0.4752 | Trait 2 |
| Reticulocyte percentage | High light scatter reticulocyte percentage | 0.8743 | 0.3795 | 1.0682 | both |
| Reticulocyte count | High light scatter reticulocyte count | 0.8694 | 7.6092 | 22.6082 | Trait 1 |
| Reticulocyte percentage | High light scatter reticulocyte count | 0.8604 | 0.3795 | 22.6082 | Trait 1 |
| Leukocyte count | Lymphocyte count | 0.8391 | 0.1494 | 0.2371 | Trait 1 |
| Reticulocyte count | High light scatter reticulocyte percentage | 0.8244 | 7.6092 | 1.0682 | both |
| Immature reticulocyte fraction | High light scatter reticulocyte percentage | 0.7328 | 3.6482 | 1.0682 | Trait 2 |
| Immature reticulocyte fraction | High light scatter reticulocyte count | 0.7106 | 3.6482 | 22.6082 | Trait 1 |
| Glucose | Glycated haemoglobin (HbA1c) | 0.6706 | 0.1372 | 0.0361 | Trait 2 |
| Leukocyte count | Neutrophil count | 0.5886 | 0.1494 | 0.1708 | Trait 1 |

**SI Tab. 3.** F values and their probabilities Pr(>F) values of traits determined in drop-one analysis. Significance thresholds are indicated by asterisks, where three asterisks indicate *p*-values below 0.001/14, two indicate p-values below 0.01/14, and one asterisk indicates *p*-values below 0.05/14.

| Trait | F value | Pr(>F) |
| --- | --- | --- |
| **BMI** | 0.741657 | 0.3892078 |
| **High light scatter reticulocyte count** | 1.761507 | 0.1845500 |
| **Erythrocyte distribution width** | 1.984917 | 0.1589896 |
| **Neutrophil count** | 9.562278 | 0.0020067 * |
| **Lymphocyte count** | 7.840082 | 0.0051467 |
| **Alkaline phosphatase** | 1.275999 | 0.2587456 |
| **Apolipoprotein A** | 2.111558 | 0.1463078 |
| **C-reactive protein** | 0.061001 | 0.8049391 |
| **Cystatin C** | 3.574881 | 0.0587677 |
| **Gamma glutamyltransferase** | 2.696038 | 0.1007155 |
| **Glucose** | 7.641474 | 0.0057432 |
| **SHBG** | 0.015972 | 0.8994413 |
| **triglycerides** | 4.532300 | 0.0333520 |
| **Vitamin D** | 8.091378 | 0.0044815 |

**SI Tab. 4.** Neutrophil cell count [10^9^ cells / liter] across cases and controls in the propensity score deciles.

SI Table 4 **SI Tab. 4.** Neutrophil cell count [ $10^9$ cells / liter] across cases and controls in the propensity score deciles.
|  | 1 | 2 | 3 | 4 | 5 | 6 | 7 | 8 | 9 | 10 |
| --- | --- | --- | --- | --- | --- | --- | --- | --- | --- | --- |
| ctrls | 3.70 | 3.83 | 4.18 | 3.98 | 4.29 | 4.48 | 4.50 | 4.79 | 4.88 | 5.23 |
| cases | 3.98 | 4.20 | 4.26 | 4.57 | 4.60 | 4.53 | 4.70 | 4.85 | 4.92 | 5.55 |

**SI Tab. 5.**
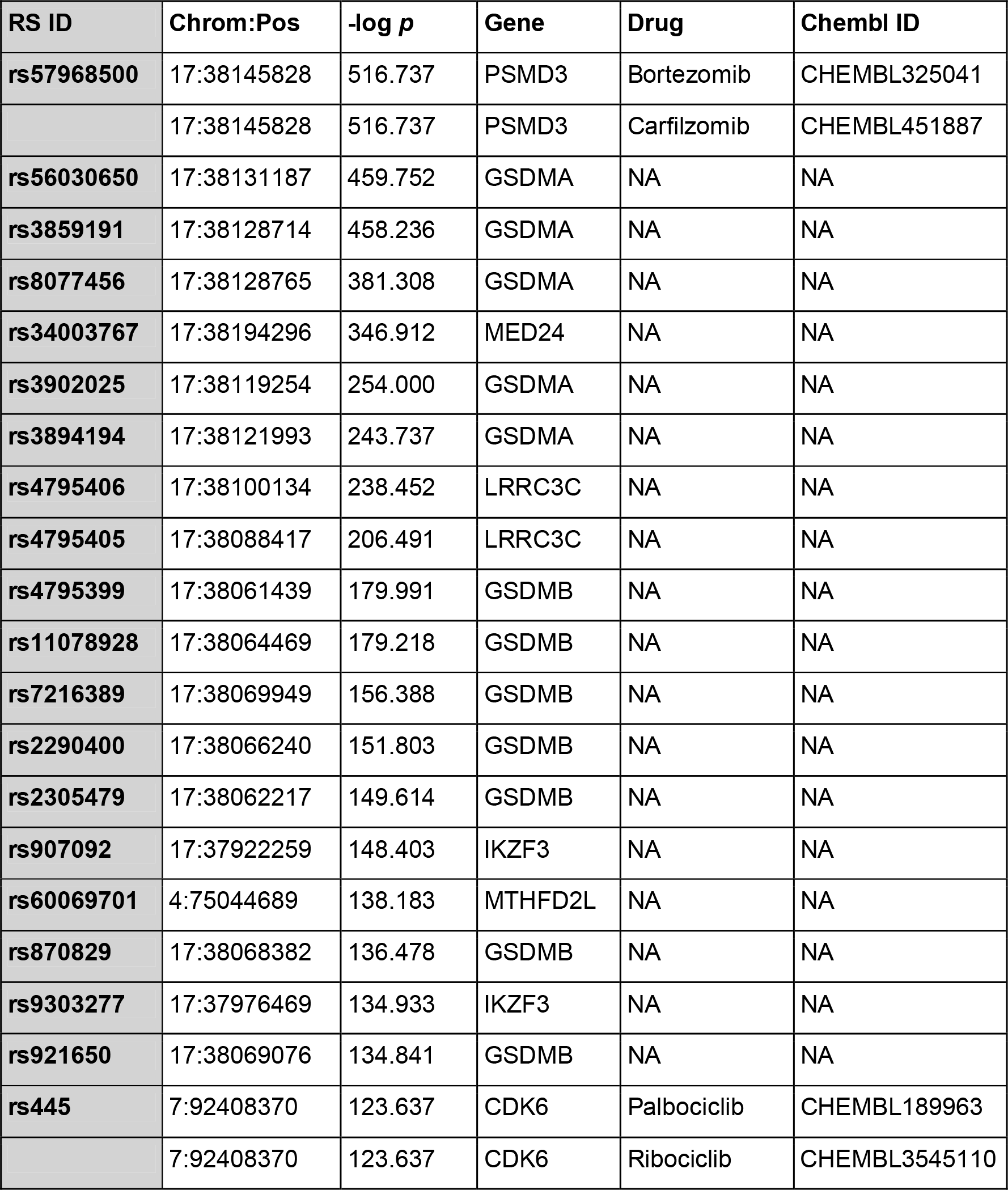
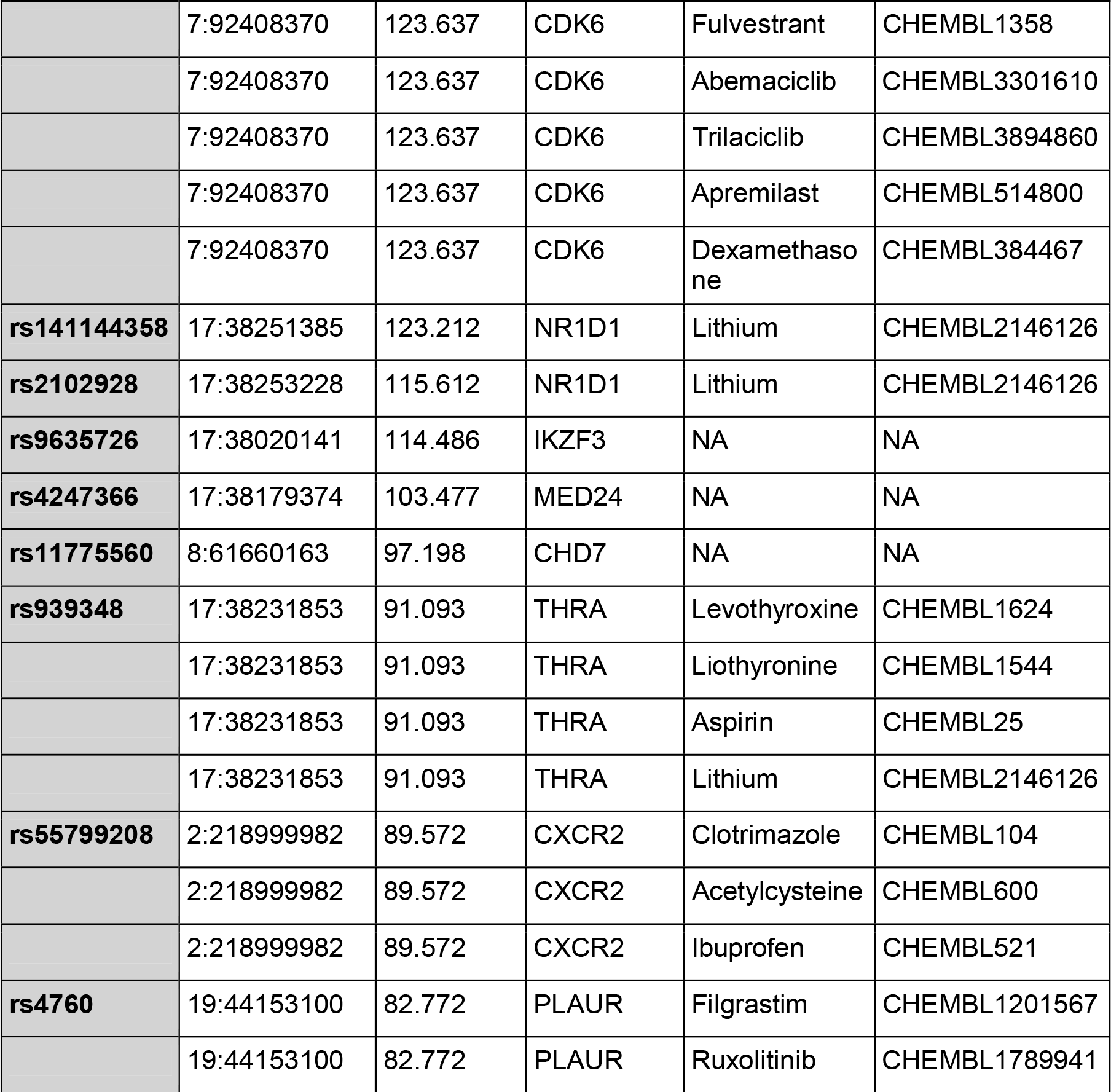
Genes and FDA-approved drugs for variants with -log *p-*values greater than 80 for neutrophil cell count. Since no clear drug-to-gene assignment was possible for the gene variants of the HLA haplotype on chromosome 6, we focused on all other significant gene variants in the following.

| RS ID | Chrom:Pos | $-\log p$ | Gene | Drug | ChEMBL ID |
| --- | --- | --- | --- | --- | --- |
| rs57968500 | 17:38145828 | 516.737 | PSMD3 | Bortezomib | CHEMBL325041 |
|  | 17:38145828 | 516.737 | PSMD3 | Carfilzomib | CHEMBL451887 |
| rs56030650 | 17:38131187 | 459.752 | GSDMA | NA | NA |
| rs3859191 | 17:38128714 | 458.236 | GSDMA | NA | NA |
| rs8077456 | 17:38128765 | 381.308 | GSDMA | NA | NA |
| rs34003767 | 17:38194296 | 346.912 | MED24 | NA | NA |
| rs3902025 | 17:38119254 | 254.000 | GSDMA | NA | NA |
| rs3894194 | 17:38121993 | 243.737 | GSDMA | NA | NA |
| rs4795406 | 17:38100134 | 238.452 | LRRC3C | NA | NA |
| rs4795405 | 17:38088417 | 206.491 | LRRC3C | NA | NA |
| rs4795399 | 17:38061439 | 179.991 | GSDMB | NA | NA |
| rs11078928 | 17:38064469 | 179.218 | GSDMB | NA | NA |
| rs7216389 | 17:38069949 | 156.388 | GSDMB | NA | NA |
| rs2290400 | 17:38066240 | 151.803 | GSDMB | NA | NA |
| rs2305479 | 17:38062217 | 149.614 | GSDMB | NA | NA |
| rs907092 | 17:37922259 | 148.403 | IKZF3 | NA | NA |
| rs60069701 | 4:75044689 | 138.183 | MTHFD2L | NA | NA |
| rs870829 | 17:38068382 | 136.478 | GSDMB | NA | NA |
| rs9303277 | 17:37976469 | 134.933 | IKZF3 | NA | NA |
| rs921650 | 17:38069076 | 134.841 | GSDMB | NA | NA |
| rs445 | 7:92408370 | 123.637 | CDK6 | Palbociclib | CHEMBL189963 |
|  | 7:92408370 | 123.637 | CDK6 | Ribociclib | CHEMBL3545110 |
|  | 7:92408370 | 123.637 | CDK6 | Fulvestrant | CHEMBL1358 |
|  | 7:92408370 | 123.637 | CDK6 | Abemaciclib | CHEMBL3301610 |
|  | 7:92408370 | 123.637 | CDK6 | Trilaciclib | CHEMBL3894860 |
|  | 7:92408370 | 123.637 | CDK6 | Apremilast | CHEMBL514800 |
|  | 7:92408370 | 123.637 | CDK6 | Dexamethasone | CHEMBL384467 |
| **rs141144358** | 17:38251385 | 123.212 | NR1D1 | Lithium | CHEMBL2146126 |
| **rs2102928** | 17:38253228 | 115.612 | NR1D1 | Lithium | CHEMBL2146126 |
| **rs9635726** | 17:38020141 | 114.486 | IKZF3 | NA | NA |
| **rs4247366** | 17:38179374 | 103.477 | MED24 | NA | NA |
| **rs11775560** | 8:61660163 | 97.198 | CHD7 | NA | NA |
| **rs939348** | 17:38231853 | 91.093 | THRA | Levothyroxine | CHEMBL1624 |
|  | 17:38231853 | 91.093 | THRA | Liothyronine | CHEMBL1544 |
|  | 17:38231853 | 91.093 | THRA | Aspirin | CHEMBL25 |
|  | 17:38231853 | 91.093 | THRA | Lithium | CHEMBL2146126 |
| **rs55799208** | 2:218999982 | 89.572 | CXCR2 | Clotrimazole | CHEMBL104 |
|  | 2:218999982 | 89.572 | CXCR2 | Acetylcysteine | CHEMBL600 |
|  | 2:218999982 | 89.572 | CXCR2 | Ibuprofen | CHEMBL521 |
| **rs4760** | 19:44153100 | 82.772 | PLAUR | Filgrastim | CHEMBL1201567 |
|  | 19:44153100 | 82.772 | PLAUR | Ruxolitinib | CHEMBL1789941 |

**SI Tab. 6.**
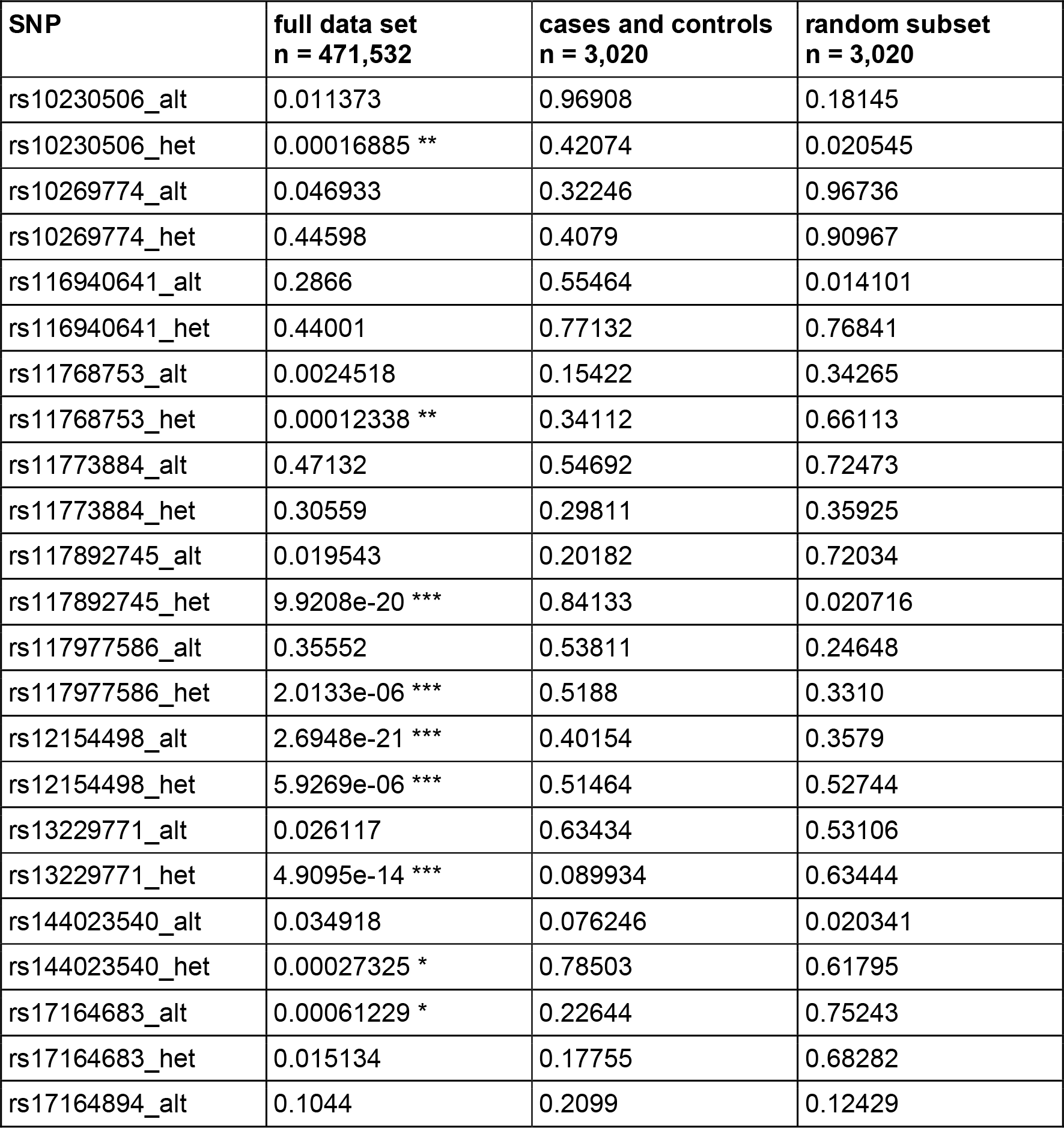
Results of the linear regressions of variants in the *CDK6* gene for neutrophil count. Suffixes indicate whether the variant is present in both alleles (“alt”) or just on one allele (“heterozygous”). Significance thresholds are indicated by asterisks, where three asterisks indicate *p*-values below 0.001/14, two indicate *p*-values below 0.01/14, and one asterisk indicates *p*-values below 0.05/14.

| SNP | full data set<br>n = 471,532 | cases and controls<br>n = 3,020 | random subset<br>n = 3,020 |
| --- | --- | --- | --- |
| rs10230506_alt | 0.011373 | 0.96908 | 0.18145 |
| rs10230506_het | 0.00016885 ** | 0.42074 | 0.020545 |
| rs10269774_alt | 0.046933 | 0.32246 | 0.96736 |
| rs10269774_het | 0.44598 | 0.4079 | 0.90967 |
| rs116940641_alt | 0.2866 | 0.55464 | 0.014101 |
| rs116940641_het | 0.44001 | 0.77132 | 0.76841 |
| rs11768753_alt | 0.0024518 | 0.15422 | 0.34265 |
| rs11768753_het | 0.00012338 ** | 0.34112 | 0.66113 |
| rs11773884_alt | 0.47132 | 0.54692 | 0.72473 |
| rs11773884_het | 0.30559 | 0.29811 | 0.35925 |
| rs117892745_alt | 0.019543 | 0.20182 | 0.72034 |
| rs117892745_het | 9.9208e-20 *** | 0.84133 | 0.020716 |
| rs117977586_alt | 0.35552 | 0.53811 | 0.24648 |
| rs117977586_het | 2.0133e-06 *** | 0.5188 | 0.3310 |
| rs12154498_alt | 2.6948e-21 *** | 0.40154 | 0.3579 |
| rs12154498_het | 5.9269e-06 *** | 0.51464 | 0.52744 |
| rs13229771_alt | 0.026117 | 0.63434 | 0.53106 |
| rs13229771_het | 4.9095e-14 *** | 0.089934 | 0.63444 |
| rs144023540_alt | 0.034918 | 0.076246 | 0.020341 |
| rs144023540_het | 0.00027325 * | 0.78503 | 0.61795 |
| rs17164683_alt | 0.00061229 * | 0.22644 | 0.75243 |
| rs17164683_het | 0.015134 | 0.17755 | 0.68282 |
| rs17164894_alt | 0.1044 | 0.2099 | 0.12429 |

**SI Tab. 7.** The two sample MR analyses here showed that for neutrophil cell count as exposure and critically ill COVID-19 status as outcome no significant effect was detected while using strict clumping parameters.

| Clumping | SNPs | beta | SE | IVW *p*-value | Pleiotropy test |
| --- | --- | --- | --- | --- | --- |
| lenient ($r = 0.2$) | 1,581 | -0.11139 | 0.04433 | 0.01199* | negative |
| strict ($r = 0.01$) | 567 | 0.01135 | 0.06987 | 0.87095 | negative |

## Notes

### Author Declarations

The research has been conducted using the UK Biobank Resource and has been approved by the UK Biobank under Application no. 36226.

### Summary of Updates

In a former version a detailed analysis of the trait genetics was missing.

